# Evaluation of nine commercial SARS-CoV-2 immunoassays

**DOI:** 10.1101/2020.04.09.20056325

**Authors:** Ria Lassaunière, Anders Frische, Zitta B. Harboe, Alex C.Y. Nielsen, Anders Fomsgaard, Karen A. Krogfelt, Charlotte S. Jørgensen

## Abstract

Due to urgency and demand, numerous severe acute respiratory syndrome coronavirus 2 (SARS-CoV-2) immunoassays are rapidly being developed and placed on the market with limited validation on clinical samples. Thorough validation of serological tests are required to facilitate their use in the accurate diagnosis of SARS-CoV-2 infection, confirmation of molecular results, contact tracing, and epidemiological studies. This study evaluated the sensitivity and specificity of nine commercially available serological tests. These included three enzyme-linked immunosorbent assays (ELISAs) and six point-of-care (POC) lateral flow tests. The assays were validated using serum samples from: i) SARS-CoV-2 PCR-positive patients with a documented first day of disease; ii) archived sera obtained from healthy individuals before the emergence of SARS-CoV-2 in China; iii) sera from patients with acute viral respiratory tract infections caused by other coronaviruses or non-coronaviruses; and iv) sera from patients positive for dengue virus, cytomegalovirus and Epstein Barr virus. The results showed 100% specificity for the Wantai SARS-CoV-2 Total Antibody ELISA, 93% for the Euroimmun IgA ELISA, and 96% for the Euroimmun IgG ELISA with sensitivities of 90%, 90%, and 65%, respectively. The overall performance of the POC tests according to manufacturer were in the rank order of AutoBio Diagnostics > Dynamiker Biotechnology = CTK Biotech > Artron Laboratories > Acro Biotech ≥ Hangzhou Alltest Biotech. Overall, these findings will facilitate selection of serological assays for the detection SARS-CoV-2-specific antibodies towards diagnosis as well as sero-epidemiological and vaccine development studies.

## Introduction

In December 2019, a novel coronavirus causing severe acute respiratory symptoms emerged in Wuhan, China [1]. The World Health Organization (WHO) termed the disease, coronavirus disease 2019 (COVID-19), and the causative virus severe acute respiratory syndrome coronavirus 2 (SARS-CoV-2). As of 7 April, the virus has spread to 212 countries and territories with 1 279 722 confirmed cases and 72 614 deaths worldwide [2]. At present, the epidemic within the majority of countries have not yet reached its peak with the number of cases and deaths predicted to rise in the coming weeks and months.

Accurate diagnosis of COVID-19 is essential, not only to ensure appropriate patient care but also to facilitate identification of SARS-CoV-2 infected people, including asymptomatic carriers, who need to be isolated to limit virus spread. The WHO recommends nucleic acid detection of SARS-CoV-2 in respiratory samples for the diagnosis of COVID-19. Unfortunately, in the face of the rapidly growing epidemics worldwide, an increased demand for diagnostic tests has led to a critical shortage in operational material for respiratory sample collection and within the molecular diagnostic workflow [3,4]. This impedes rapid large scale testing, a necessity for controlling the epidemic. Moreover, the heterogeneity of respiratory sample material and anatomical location of sample collection, for example throat swab, saliva or endotracheal aspirate, affect the sensitivity of SARS-CoV-2 viral nucleic acid testing [5,6]. Overall, there is an urgent need to identify alternative diagnostic means.

Antibody testing, either using enzyme-linked immunosorbent assay (ELISA) or point-of-care (POC) lateral flow immunoassays, may overcome some of these challenges. SARS-CoV-2-specific antibodies can be detected in in serum of approximately 40% of COVID-19 patients as early as seven days after the onset of symptoms, with seroconversion rates rapidly increasing to >90% by day 14 [7]. In recent studies, antibody testing has been shown to be more sensitive than viral nucleic acid detection after approximately eight days of COVID-19 illness duration [7,8]. While the combination of PCR and antibody tests is optimal for accurate diagnosis [6], antibody detection will be particularly relevant for the later stages of infection where the virus has been eliminated [5]. In addition to the diagnostic value of antibody testing, it will identify individuals who developed immunity after infection that may protect against subsequent re-infection [9], as well as define and monitor the extent of virus spread and a population’s herd immunity on a societal level.

Antibody testing may therefore be relevant in the following settings: i) diagnosis of patients who seek medical attention more than seven after the onset of symptoms; ii) contact tracing; iii) determining potential immunity and risk of infection; and iv) sero-epidemiological studies to understand the extent of COVID-19 spread. Due to urgency and demand, a lot of serological tests are rapidly being developed and made available on the market with only limited validation on clinical samples. To address this, the present study evaluated three ELISA assays and six POC lateral flow immunoassays for the detection of SARS-CoV-2 antibodies in patients with COVID-19. CE-marked SARS-CoV-2 ELISA assays and POC tests were selected based on availability in Denmark at the time of testing.

## Materials & Methods

### Study design

Retrospective study evaluating the sensitivity and specificity of commercially available immunoassays for the detection of antibodies specific to the SARS-CoV-2 virus. Patient serum samples used in this study were submitted to the routine serology laboratory at Statens Serum Institut for diagnostic purposes.

### Serum samples

Case serum samples were obtained from COVID-19 patients (n = 30) admitted to the intensive care unit at Hillerød Hospital, Denmark. SARS-CoV-2 infection was confirmed by viral nucleic acid detection in samples from the respiratory tract. Control serum samples included archived anonymous serum samples obtained from healthy blood donors 18-64 years with no history of SARS-CoV-2 infection (n = 10) and no recent travel history, sera from patients with acute viral respiratory tract infections caused by other coronaviruses (n = 5) or non-coronaviruses (n = 45), and sera from patients positive for dengue virus (n = 9), cytomegalovirus (CMV; n = 2) and Epstein Barr virus (EBV; n = 10). One patient was positive for both CMV and EBV.

### ELISA assays

The Wantai SARS-CoV-2 Ab ELISA (Beijing Wantai Biological Pharmacy Enterprise, Beijing, China; Cat # WS-1096) was performed according to the manufacturer’s instructions. The assay is based on a double-antigen sandwich principle that detects total antibodies binding SARS-CoV-2 spike protein receptor binding domain (RBD) in human serum or plasma. Briefly, 100 μl undiluted serum samples were added to wells coated with recombinant SARS-CoV-2 antigen and incubated for 30 minutes at 37 °C. Wells were washed five times followed by the addition of HRP-conjugated SARS-CoV-2 antigen and subsequent incubation for 30 minutes at 37 °C. Wells were washed five times and a chromogen solution was added. Following 15 minutes of incubation at 37 °C, the reaction was stopped and the resultant absorbance was read on a microplate reader at 450 nm with reference at 620 nm. The cut-off value for a positive result was calculated according to the manufacturer’s instruction by adding the calculated negative control value to 0.160.

The Anti-SARS-CoV-2 IgG and IgA ELISAs (Euroimmun Medizinische Labordiagnostika, Lübeck, Germany; Cat # EI 2668-9601 G and EI 2606-9601 A, respectively) were performed according to the manufacturer’s instructions. In two separate semi-quantitative ELISAs, either IgA or IgG antibodies against SARS-CoV-2 spike protein subunit 1 (S1) are detected in human serum or plasma. Briefly, 1:101 diluted serum samples were added to wells coated with recombinant SARS-CoV-2 antigen and incubated for 60 minutes at 37 °C. Wells were washed three times followed by the addition of HRP-conjugated anti-human IgA or IgG and subsequent incubation for 30 minutes at 37 °C. Wells were washed three times and a chromogen solution was added. Following 30 minutes of incubation at room temperature, the reaction was stopped and the resultant absorbance was read on a microplate reader at 450 nm with reference at 620 nm. A ratio between the extinction of the sample and calibrator on each plate were calculated. According to the manufacturer’s recommendations, a ratio <0.8 is considered negative, ≥0.8 and <1.1 borderline, and ≥1.1 positive. However, for sensitivity and specificity, 1.1 was used as a more stringent cut-off value for positive results and all values <1.1 were considered negative.

### Point-of-care (POC) tests

Six POC tests for rapid detection of antibodies in blood, serum or plasma were evaluated: 2019-nCOV IgG/IgM Rapid Test (Dynamiker Biotechnology, Tianjin, China Cat # DNK-1419-1), OnSiteTM COVID-19 IgG/IgM Rapid Test (CTK Biotech, Poway, CA, USA; Cat # R0180C), Anti-SARS-CoV-2 Rapid Test (AutoBio Diagnostics, Zhengzhou, China; Cat # RTA0204), Coronavirus Diseases 2019 (COVID-19) IgM/IgG Antibody Test (Artron Laboratories, Burnaby, Canada; Cat # A03-51-322), 2019-nCoV IgG/IgM Rapid Test Cassette (Acro Biotech, Rancho Cucamonga, CA, USA; Cat # INCP-402), and 2019-nCoV IgG/IgM Rapid Test Cassette (Hangzhou Alltest Biotech, Hangzhou, China; Cat # INCP-402). The tests were performed at room temperature according to the manufacturer’s instructions. For all tests, the recommended sample volume of 10 μl serum was added to the specimen well on the individual test cassettes followed by the addition of the supplied buffer. The buffer volume differed by manufacturer and was added accordingly (60 μl, three drops, two drops, two drops, two drops and 60 μl, respectively). The result was read visually after 10 minutes. Weak signals for IgM and IgG, together or separate, was considered positive.

### Statistical analyses

Sensitivity was defined as the proportion of patients correctly identified as having SARS-CoV-2 infections, as initially diagnosed using nucleic acid detection of SARS-CoV-2 in respiratory samples. Specificity was defined as the proportion of SARS-CoV-2 immune naïve study participants accurately identified as negative for COVID-19. The clinical accuracies of the ELISA assays were examined by using Receiver Operator Characteristic (ROC) plots with GraphPad Prism version 8.0.2 (GraphPad Software, San Diego, CA, USA). ROC area under the curve (AUC) were calculated as the fraction “correctly identified to be positive” and the fraction “falsely identified to be positive” determined according to manufacturer cut-off values for positive results.

## Results

### Sensitivities and specificities of the ELISA assays

Three commercial CE-marked ELISA assays for detecting SARS-CoV-2 antibodies were evaluated using 30 serum samples from PCR-positive cases with SARS-CoV-2 and 82 control serum samples. Twenty nine of the 30 cases (97%) were positive for SARS-CoV-2-specific antibodies by at least one of the three ELISA assays. In one case, only a positive IgA result was detected, while in another case antibody responses were negative by all tests.

The distance of data points from the manufacturer recommended cut-off values and confidence in assigning a positive or negative status differed between the three assays (Figure 1A). The distribution of positive and negative data points were distinct for the Wantai Total Ab assay, with a cut-off value above all the control sera samples, which allowed for unequivocal interpretation. Conversely, the Euroimmun IgA and IgG assays data had a less distinct separation, resulting in a ‘grey zone’ of borderline data points to which a positive or negative status could not be assigned. Both case and control sera had borderline or inconclusive data points.

**Figure 1.**
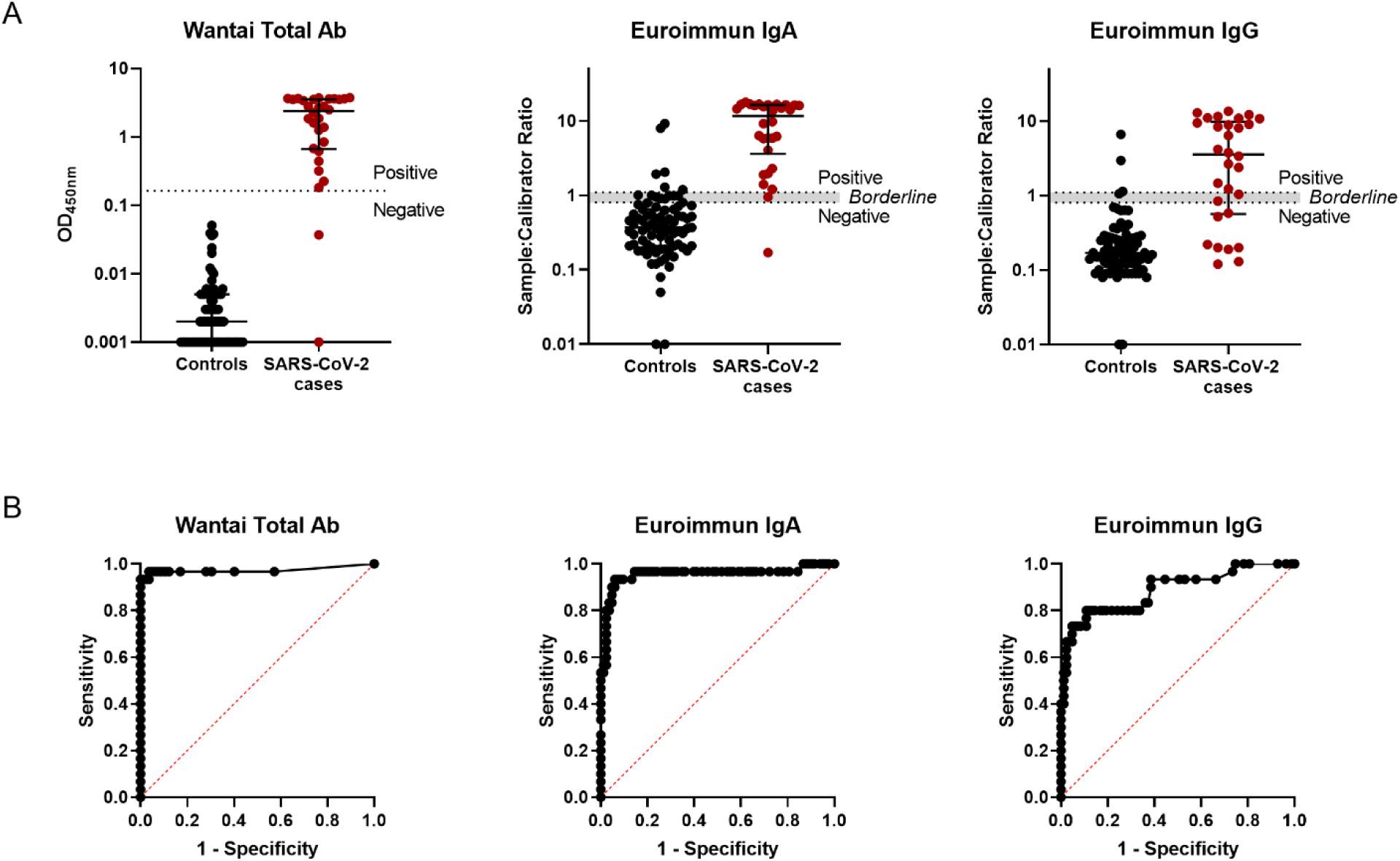
SARS-CoV-2 antibody ELISA assay performance. A) ELISA data distribution obtained for control and SARS-CoV-2 case sera using three commercial ELISA kits. Lines represents median values with interquartile ranges. The dotted lines indicate the respective cut-off values recommended by manufacturer to determine positive and negative test results. The grey zone marks the ratio range with borderline results. B) ROC curves for the respective ELISAS assays.

The sensitivities and specificities are shown in table 1. The sensitivity of the Wantai Total Ab ELISA was equivalent to that of Euroimmun’s IgA ELISA at 93% and was greater than that observed for Euroimmun’s IgG ELISA at 67%. The specificity of the Wantai Total Ab ELISA was 100% compared to 93% and 96% for the Euroimmun IgA and IgG ELISAs, respectively. The positive predictive value and negative predictive value was the highest for the Wantai Total Ab ELISA at 100% and 98%, respectively, compared to the Euroimmun IgA ELISA (82% and 97%, respectively) and IgG ELISA (87% and 89%, respectively). The Euroimmun IgA ELISA cross-reacted primarily with serum that contained antibodies to more than one respiratory virus (4/6 [67%]) and associated with the presence of adenovirus antibodies (5/6 [83%]) and dengue virus antibodies (Table 2). The Euroimmun IgG ELISA cross-reacted with a serum sample positive for human coronavirus HKU1 and two samples with adenovirus antibodies.

**TABLE 1.**
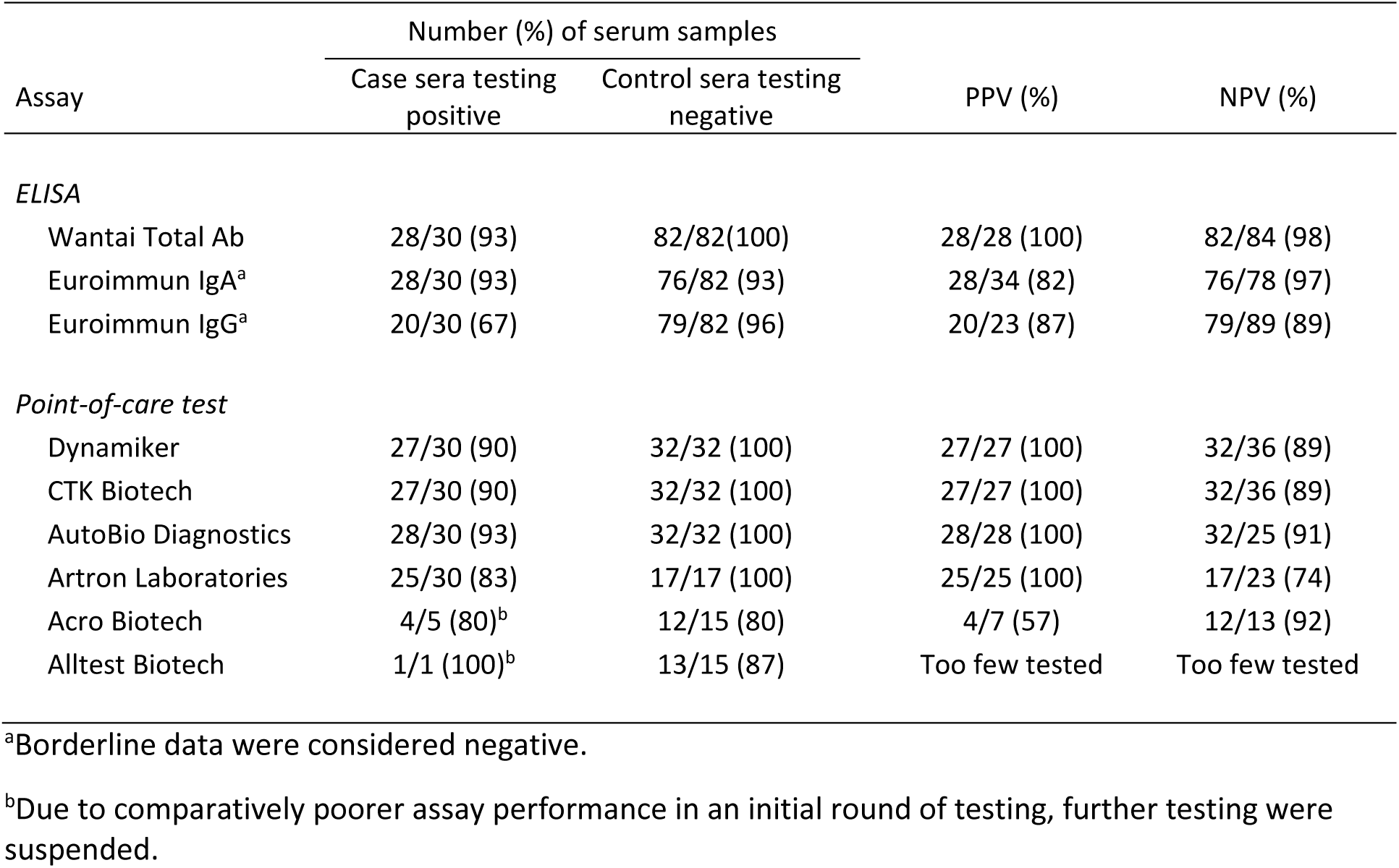
Analytical sensitivities, specificities, and predictive values for SARS-CoV-2 antibody detection

| Assay | Number (%) of serum samples |  | PPV (%) | NPV (%) |
| --- | --- | --- | --- | --- |
|  | Case sera testing positive | Control sera testing negative |  |  |
| *ELISA* |  |  |  |  |
| Wantai Total Ab | 28/30 (93) | 82/82(100) | 28/28 (100) | 82/84 (98) |
| Euroimmun IgA<sup>a</sup> | 28/30 (93) | 76/82 (93) | 28/34 (82) | 76/78 (97) |
| Euroimmun IgG<sup>a</sup> | 20/30 (67) | 79/82 (96) | 20/23 (87) | 79/89 (89) |
| *Point-of-care test* |  |  |  |  |
| Dynamiker | 27/30 (90) | 32/32 (100) | 27/27 (100) | 32/36 (89) |
| CTK Biotech | 27/30 (90) | 32/32 (100) | 27/27 (100) | 32/36 (89) |
| AutoBio Diagnostics | 28/30 (93) | 32/32 (100) | 28/28 (100) | 32/25 (91) |
| Artron Laboratories | 25/30 (83) | 17/17 (100) | 25/25 (100) | 17/23 (74) |
| Acro Biotech | 4/5 (80)<sup>b</sup> | 12/15 (80) | 4/7 (57) | 12/13 (92) |
| Alltest Biotech | 1/1 (100)<sup>b</sup> | 13/15 (87) | Too few tested | Too few tested |

<sup>a</sup>Borderline data were considered negative.
<sup>b</sup>Due to comparatively poorer assay performance in an initial round of testing, further testing were suspended.

**TABLE 2.** False positive results for sera from controls diagnosed with infections other than SARS-CoV-2

| Virus-specific antibodies detected | ELISA |  |  |  |  | Point-of-Care Tests |  |  |
| --- | --- | --- | --- | --- | --- | --- | --- | --- |
|  | # sera tested | Euroimmun IgA |  | Euroimmun IgG |  | # sera tested | Acro Biotech | Alltest Biotech |
|  |  | Positive | Borderline | Positive | Borderline |  |  |  |
| Unknown | 10 |  | 1 |  | 1 | 2 |  |  |
| hCoV-OC43 | 1 |  |  |  |  | 1 |  |  |
| hCoV-HKU1 | 2 |  |  | 1 |  | 2 | 1 (IgM only) |  |
| hCoV-NL63 | 2 |  |  |  |  | 2 |  |  |
| Adv | 4 | 1 | 1 | 1 |  | 1 |  |  |
| Infl A | 5 |  |  |  |  | 0 |  |  |
| Infl B | 7 |  |  |  |  | 0 |  |  |
| Infl A, Infl B | 9 |  | 2 |  |  | 1 |  |  |
| Infl A, Infl B, Adv | 6 | 1 | 2 | 1 |  | 3 | 1 (IgM only) | 1 (IgM only) |
| Infl A, Adv | 4 | 1 |  |  |  | 1 |  |  |
| Infl B, Adv | 2 |  |  |  |  | 1 |  |  |
| Infl A, Infl B, RSV | 2 |  |  |  |  | 0 |  |  |
| Infl A, Infl B, RSV, Adv | 3 |  |  |  |  | 2 |  |  |
| Adv, RSV | 1 | 1 |  |  |  | 1 |  |  |
| Infl A, RSV, Adv | 1 | 1 |  |  |  | 1 |  |  |
| Infl B, RSV | 1 |  |  |  |  | 0 |  |  |
| EBV | 10 |  | 2 |  |  | 0 |  |  |
| CMV | 1 |  |  |  |  | 0 |  |  |
| CMV, EBV | 2 |  |  |  |  | 0 |  |  |
| Dengue | 9 | 1 | 1 |  |  | 2 | 1 (IgM only) | 1 (IgM only) |
| Total | 82 | 6 | 9 | 3 | 1 | 20 | 3 | 2 |

hCoV – human coronavirus; Adv – adenovirus; Infl – influenza; RSV – respiratory syncytial virus; EBV – Epstein-Barr virus; CMV – cytomegalovirus.

### ROC analysis

ROC AUC analysis presents a good parameter for the diagnostic power of an individual test and were compared between the different ELISA kits (Figure 1B). The Wantai total antibody kit had the highest measure at 0.973 (95% CI: 0.921-1.000), followed by Euroimmun’s IgA ELISA with 0.954 (95% CI: 0.897-1.00) and Euroimmun’s IgG ELISA with 0.887 (95% CI: 0.810-0.964).

### Sensitivities and specificities of the POC tests

Four POC tests were tested on all 30 case serum samples and had sensitivities in the rank order of 93% for AutoBio Diagnostics, 90% Dynamiker Biotechnology and CTK Biotech, and 83% for Artron Laboratories (Table 1). The positive predictive value of these tests were 100%, while the negative predictive values were 91%, 89%, 89%, and 74%, respectively. The Acro Biotech was evaluated on five case serum samples and had a specificity of 80%. One case serum sample was tested with the Hangzhou AllTest Biotech test and was positive for both IgM and IgG.

The specificity of the six POC tests were evaluated primarily on control samples that showed some cross-reactivity in the SARS-CoV-2 ELISA assays; the number of control sera tested varied between the different POC tests (Table 1). The POC tests manufactured by Dynamiker Biotechnology, CTK Biotech, AutoBio Diagnostics and Artron Laboratories had a 100% specificity, whereas the test from Acro Biotech and Hangzhou AllTest Biotech had a specificity of 80% and 87%, respectively. For the latter two tests, cross-reactivity was only observed for IgM. The Acro Biotech test cross-reacted with a control serum sample from a human coronavirus HKU1 patient.

### Antibody detection relative to duration of illness

To evaluate the sensitivities of the assays at different stages of COVID-19 disease, case sera were grouped according the duration of disease: early phase, 7 to 13 days after the onset of disease symptoms; middle phase, 14 to 20 days after the onset of disease symptoms; and late phase, ≥21 days after the onset of disease symptoms. The sensitivities of the assays ranged from 40 to 86% for the early phase samples, 67 to 100% for the middle phase samples, and 78 to 89% for the late phase (Figure 2).

**Figure 2.**
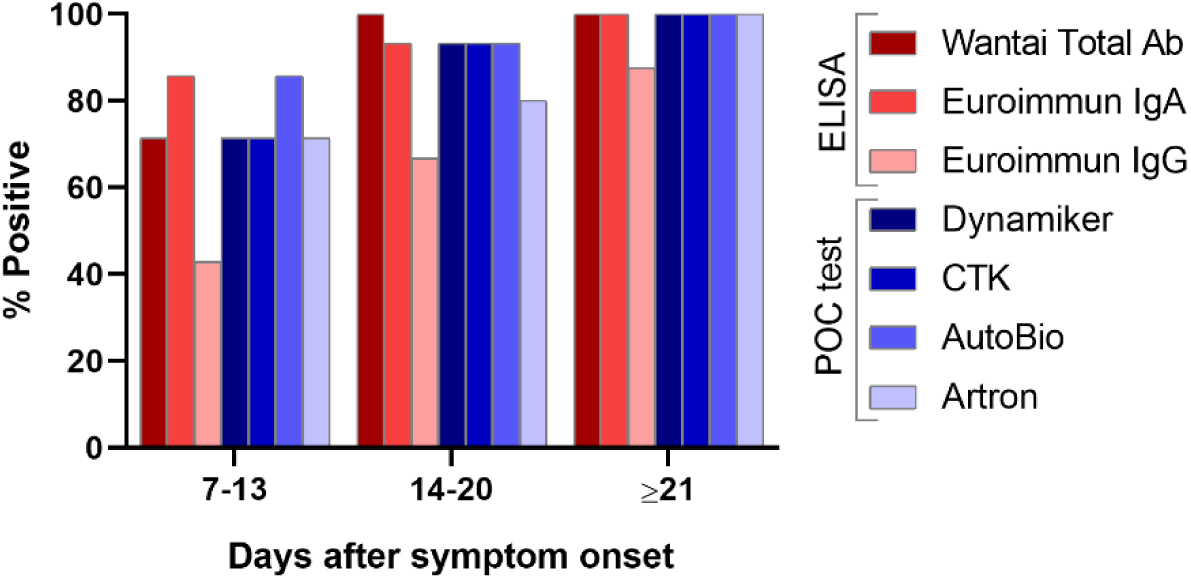
Analytical sensitivities of SARS-CoV-2 serologic assays in relation to the duration of illness: 7 to 13 days (n = 7), 14 to 20 days (n = 15), and ≥21 days (n = 8).

In the early phase, the Wantai Total Ab ELISA had a sensitivity of 71% that plateaued at 100% after 10 days of illness duration. The IgG ELISA had the lowest sensitivity at all three phases that showed a distinct increase with each consecutive phase i.e. 43% in the early phase, 67% in the middle phase, and 78% in the late phase. While the four POC tests evaluated according to illness duration were often weakly positive or detected only IgG or IgM during the early phase (data not shown), their sensitivities were comparable to the Wantai Total Ab ELISA and Euroimmun IgA ELISA in all three phases. In the early phase, a case sample that was negative by both Total Ab and IgG was positive in the IgA ELISA.

### Agreement between serological assays

To determine the agreement between the different ELISAs and POC tests evaluate, the proportion of case sera that shared the same result between two assays were calculated. Despite comparable sensitivities of certain assays, the tests did not necessarily give the same result in all instances (Figure 3A). The only tests that were 100% concordant were the Dynamiker Biotechnology and CTK Biotech POC tests (Figure 3B). The Wantai Total Ab ELISA and Euroimmune IgA ELISA was 93% concordant, whereas the highest agreement between an ELISA and POC test was 93% between the Wantai Total Ab ELISA and AutoBio Diagnostics POC test.

**Figure 3.**
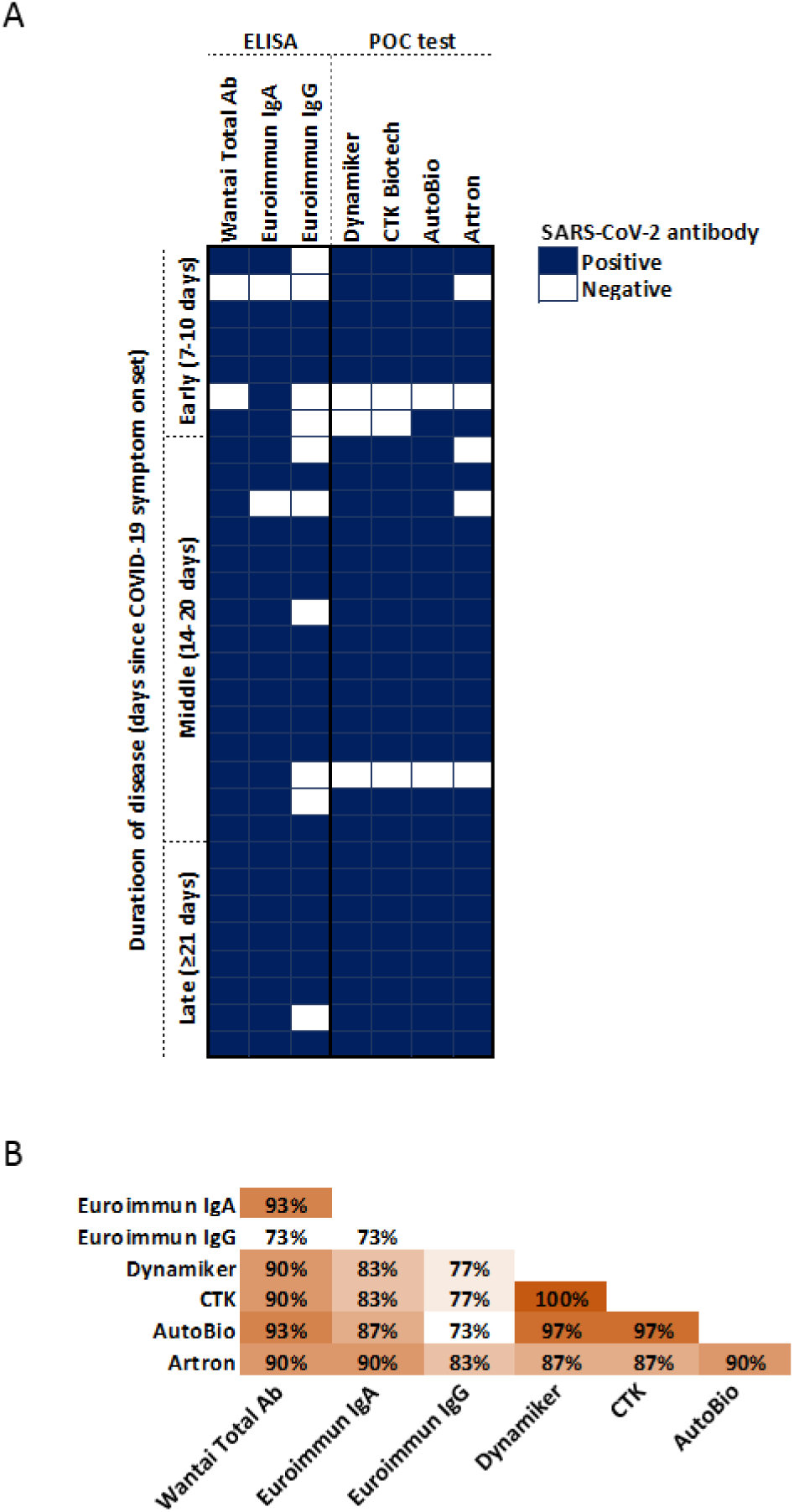
Agreement between ELISA and POC test results. A) Individual COVID-19 patient with PCR-confirmed SARS-CoV-2 infection. Each row represents a patient, each column a serological test, a blue block represents a positive result and a white block a negative result. B) The proportion of results that agreed between two assays.

## Discussion

In the present study, three SARS-CoV-2-specific commercial ELISA assays and six POC rapid tests were evaluated using sera from hospitalized adult patients with PCR-confirmed diagnoses for SARS-CoV-2 and a collection of control serum samples taken before the emergence of the virus in China in December 2019. Overall, the Wantai Total Ab ELISA had superior sensitivity and specificity compared to both Euroimmun IgA and IgG ELISAs. The POC tests varied notably, with the best performance observed for the test produced by AutoBio Diagnostics, followed by the tests produced by Dynamiker Biotechnology and CTK Biotech.

The differences observed for the sensitivity and specificity of the SARS-CoV-2-specific total antibody testing and antibody type ELISAs correspond to previous reports. The Wantai Total Ab ELISA performed as reported by the manufacturer (94.5% sensitivity and 100% specificity) and a separate study (93.1% sensitivity) where the Total Ab ELISA, IgM ELISA and IgG ELISA produced by Beijing Wantai Biological Pharmacy Enterprise were compared [7]. In the latter study, the IgM and IgG ELISAs had lower sensitivities (83% and 65%, respectively) compared to the Total Ab ELISA (93.1%) [7]. This notably lower sensitivity for SARS-CoV-2-specific IgG detection is in agreement to that observed here for the Euroimmun IgG ELISA (65%). In the present study the Wantai IgG and Euroimmun IgG ELISAs could not be compared due to unavailability of the former. The possibility that overall lower sensitivity of SARS-CoV-2 IgG ELISAs may be a more universal occurrence rather than manufacturer dependent warrants further investigation. In addition to lower sensitivities, the Euroimmun IgA and IgG ELISAs are also more prone to cross-react with negative sera as described in the present study and in a separate analysis of the beta-versions of these assays [10].

The differences between the assays may, in part, be explained by the SARS-CoV-2 antigen targeted and the ELISA format used. Both kits detect antibodies to the S1 subunit of the SARS-CoV-2 spike protein; however, the Wantai Total Ab ELISA only targets the RBD within the S1. The RBD represents approximately 33% of the S1 subunit, thus epitopes that may be recognized by cross-reacting epitopes outside of this domain are absent. Furthermore, the RBD is highly diverse between SARS-CoV-2 and other beta-coronaviruses (hCoV-OC43 and hCoV-HKU1), which may further reduce the likelihood of cross-reaction with these circulating coronaviruses. Moreover, the Wantai Total Ab ELISA uses an antigen-antibody-antigen(peroxidase) format whereas the Euroimmun ELISAs employ an antigen-antibody-antibody(peroxidase) format. The specificity of the former is determined by a single antibody, whereas the latter has a second antibody that may introduce additional specificities. The antigen-antibody-antibody format is required to distinguish between specific antibody types, but may not necessarily have lead to decreased specificity as shown for in-house ELISAs [10].

The clinical sensitivity of IgM for early diagnosis of COVID-19 is currently unclear. SARS-CoV-2-specific IgM does not consistently appear before its IgG counterpart, with some studies reporting detection of SARS-CoV-2 spike protein-specific IgG before IgM [6,7,11]. While all the POC tests evaluated in this study are capable of detecting both SARS-CoV-2 IgM and IgG antibodies, the majority detected both antibody types simultaneously, even in the early convalescent phase, while some detected only IgG and others only IgM (data not shown). On the contrary, cross-reactive IgM antibodies resulted in decreased specificity of two POC tests evaluated. This IgM-associated reduced specificity was not observed for all POC tests, thus other unknown factors define POC test specificity. POC tests are, by definition, often performed beside the patient, whereas ELISAs are conducted in laboratories. This study demonstrated that while certain ELISA assays and POC tests may share similar sensitivities and specificities, their results may not necessarily correspond for a single patient sample. These discrepancies should be noted where SARS-CoV-2 ELISAs are used to confirm POC test results.

Since the appearance of antibodies is time dependent, diagnosis of COVID-19 by serological methods is limited to patients with a longer duration of illness. Within seven days of symptom onset or in the acute phase of disease, nucleic acid detection of SARS-CoV-2 in respiratory samples is superior to antibody detection for the diagnosis of COVID-19 [7,8,12]. However, after eight days of illness, the sensitivity of serological assays surpasses that of nucleic acid testing [7,8]. Here we reported a 100% seropositivity in patients 10 days after the onset of symptoms. It is unclear whether the latter sensitivity can be extrapolated to mild COVID-19 cases, since the present study comprised severely ill adult COVID-19 patients only. However, the sensitivity of the assay may not necessarily be very affected, since reports show similar seroconversion rates for patients with mild and severe COVID-19 disease despite generally lower SARS-CoV-2 antibody titres in the former group [6,7,13]. Conversely, studies on the dynamics and detection of SARS-CoV-2 antibodies in children are lacking and requires urgent attention.

In conclusion, our findings show that in an ELISA format the sensitivity of detecting total SARS-CoV-2 RBD-specific antibodies is higher than that of assays detecting spike-specific IgA or IgG only. It is important to note that the presence of SARS-CoV-2-specific antibodies does not necessarily correspond to protection against SARS-CoV-2 infection and disease. In order to define antibody-mediated protection, further investigation of virus-specific antibody functions that include neutralization and Fc-mediated effector functionality are needed. Sero-epidemiological investigations together with longitudinal studies on sequential samples taken from SARS-CoV-2 patients are necessary to characterize the spread of the virus and the long term protection of the antibodies measured.

## Data Availability

The data are available from the corresponding author by request.

## Acknowledgements

The research has been conducted using the Danish National Biobank resource, supported by the Novo Nordisk Foundation, grant number 2010-11-12 and 2009-07-28.

## Ethical statement

Exemption for review by the ethical committee system and informed consent was given by the Committee on Biomedical Research Ethics - Capital Region in accordance with Danish law on assay development projects.

## Conflicts of interest

The authors declare no competing interests.

## Notes

### Competing Interest Statement

The authors have declared no competing interest.

